# Socioeconomic Inequalities in the Visual Assessment of Older Adults with Cataracts in Peru

**DOI:** 10.1101/2024.06.06.24308564

**Authors:** Claudio Intimayta-Escalante, Gino Vitteri-Soto

## Abstract

**Introduction:** In Peru, one in six adults over 50 years old has cataracts. This proportion increases due to the lack of routine visual assessments among older adults. Therefore, the objective of this research was to evaluate the socioeconomic inequalities in visual assessments among older adults with cataracts in Peru.

**Methods:** This analytical cross-sectional study was conducted using data from the Demographic and Family Health Survey in Peru from 2013 to 2023. Socioeconomic conditions such as gender, age, educational level, wealth quintile, area or place of residence, health insurance status, and comorbidities were evaluated. Inequalities were estimated using the concentration index (CI) and Erreyger’s modification (ECI).

**Results:** Of the 6,367 older adults with cataracts included in the study, 93.1% (95%CI: 92.2–93.9) had a visual assessment. The average age of the participants was 76.7 years, with over half of them being women (56.2%). Conditions such as educational level, wealth quintile, area or place of residence, and comorbidities mediated differences in the proportion of adults with cataracts who had visual assessments (p*<*0.001). Adults without education (ECI: -0.16), residents of rural areas (ECI: -0.41), or those outside Lima (ECI: -0.51) exhibited greater inequalities in receiving visual assessments.

**Conclussion:** Among older adults with cataracts, living in rural areas or outside Lima and having a low educational level were identified as key factors mediating greater inequality in visual assessments.

## INTRODUCTION

By 2020, it is estimated that nearly 94 million people will be blind or visually impaired (1). The leading cause of this problem is cataract, a condition that can be managed with early detection, timely treatment, and surgical intervention (2, 3). While aging is a common factor, poverty, lack of education, and limited access to eye care can increase the risk of cataracts (4, 5). Thus, the gap in access to these services varies in the population according to income, educational level, age, sex, place or area of residence, and even ethnic identification, leading to disparities in eye care access (6–11).

The global prevalence of visual impairment due to cataracts ranges from 0.9% to 10.7%, reflecting differences in health-care systems, socioeconomic conditions, and availability of treatment (12). In South America, disparities in healthcare infrastructure, awareness, and treatment accessibility contribute to the varying prevalence of cataract-related blindness among older adults (13, 14). In Peru, 13.5% of the population has visual impairment, and one in six adults over 50 has cataracts, showing the heavy burden of eye conditions in the country (15, 16). Aging, delayed diagnosis, and health conditions such as diabetes significantly contribute to the increased prevalence of cataracts (17–23).

Current visual health guidelines in Peru do not adequately address the socioeconomic and health disparities in access to and utilization of eye care services (24–26). These differences in eye care can negatively impact older adults, reducing their quality of life, functional independence and increasing the risk of accidents (27–31). This study aimed to assess socioeconomic disparities in evaluating the vision of older adults with cataracts in Peru, looking at differences in accessing eye care services and treatment results.

## METHODS

### Study Design

An analytical study was conducted using data from the Demographic and Family Health Survey (or ENDES, in Spanish) in Peru. The National Institute of Statistics and Informatics (or INEI, in Spanish) has been conducting the ENDES annually in Peru, a country in South America with approximately 32 million of inhabitans, wich capital city is Lima (32). Therefore, we assessed Peruvian adults aged 65 or older with cataracts who were surveyed in ENDES between 2013 and 2023.

### Variables Assessed

Within the survey, participants are asked whether they have ‘been diagnosed with cataract’ and whether a ‘doctor or health professional has evaluated or measured your vision’. In addition, other variables were assessed, such as sex (male or female), age group (65 to 69, 70 to 79, and 80 or older), education level (no education, primary, secondary, or higher), wealth quintile (first, second, third, fourth, and last quintile), wealth quintile (first, second, third, fourth, and last quintile), area of residence (rural and urban), living in the capital city (yes or no), having health insurance (yes or no), and having a comorbidity such as overweight or obesity, hypertension, or diabetes mellitus.

### Statistical Analysis

The analysis was performed using R Studio software version 4.2.2 (https://cran.r-project.org/), including the complex sample design of ENDES. Categorical variables were described using frequencies and percentages, with their respective 95% confidence intervals weighted by the design effect. In addition, the Rao-Scott test was used to assess differences in the proportion of adults with cataracts who had an ophthalmological examination between the categories of variables included in the study. We used Poisson regression models with large variances to look at the relationship between socioeconomic factors and eye exams. This helped us figure out the crude Prevalence Ratio (cPR) and the adjusted Prevalence Ratio (aPR).

### Inequality Analysis

Inequality analysis utilized concentration curves to show the disparity in eye assessments based on socioeconomic status, ranging from the poorest to the richest wealth quintile. So, the concentration index (CI) was found by guessing the area above or below the curve. This way, the conditions that led to crowding above the curve showed how poverty affected inequality (33). Additionally, the Erreygers Concentration Index (ECI) was utilized to calculate inequality, considering the extremes of the population distribution across wealth quintiles for a more equitable assessment (34). Also, maps of Peru were generated to show the inequal geographic distribution of visual assessment among adults over 64 years old, along with the ECI distribution.

The study also evaluated disparities among participants with specific comorbidities like overweight, obesity, hypertension, and diabetes mellitus. Additionally, the study analyzed annual changes in the percentage of adults with cataracts who received eye exams and examined the related inequality indices to track trends over time.

### Ethical Aspects

The study utilized data from ENDES, a national survey conducted with participants’ informed consent, for its development. In addition, the study had no information to identify the participants included in the research.

## RESULTS

Of the 6,367 older adults with cataracts examined (Appendix 1), more than half were female (56.2%, 95%CI: 53.7 to 58.7) and had a mean age of 76.7 years (95%CI: 76.3 to 77.0). In terms of socio-economic conditions, 39.9% (95%CI: 37.7 to 42.1) had completed secondary or higher education, and 56.0% (95%CI: 53.7 to 58.2) were in the first two wealth quintiles. Furthermore, approximately 84.2% (95%CI: 93.2 to 95.2) of the surveyed adults resided in urban areas, with around 47.1% (95%CI: 44.8 to 49.4) living in the capital. In addition, 87.7% (95%CI: 84.9 to 90.0) had health insurance, and 77.3% (95%CI: 75.2 to 79.9) had a comorbidity (Table 1). In addition, 64.0% (95%CI: 61.5 to 66.4) were overweight or obese, 46.5% (95%CI: 44.1 to 49.0) had high blood pressure, and 14.1% (95%CI: 10.8 to 18.3) had diabetes mellitus. Among older adults with cataracts, only six in 100 had no visual examination or measurement (6.9%; 95%CI: 6.1 to 7.8).

**Table 1.**
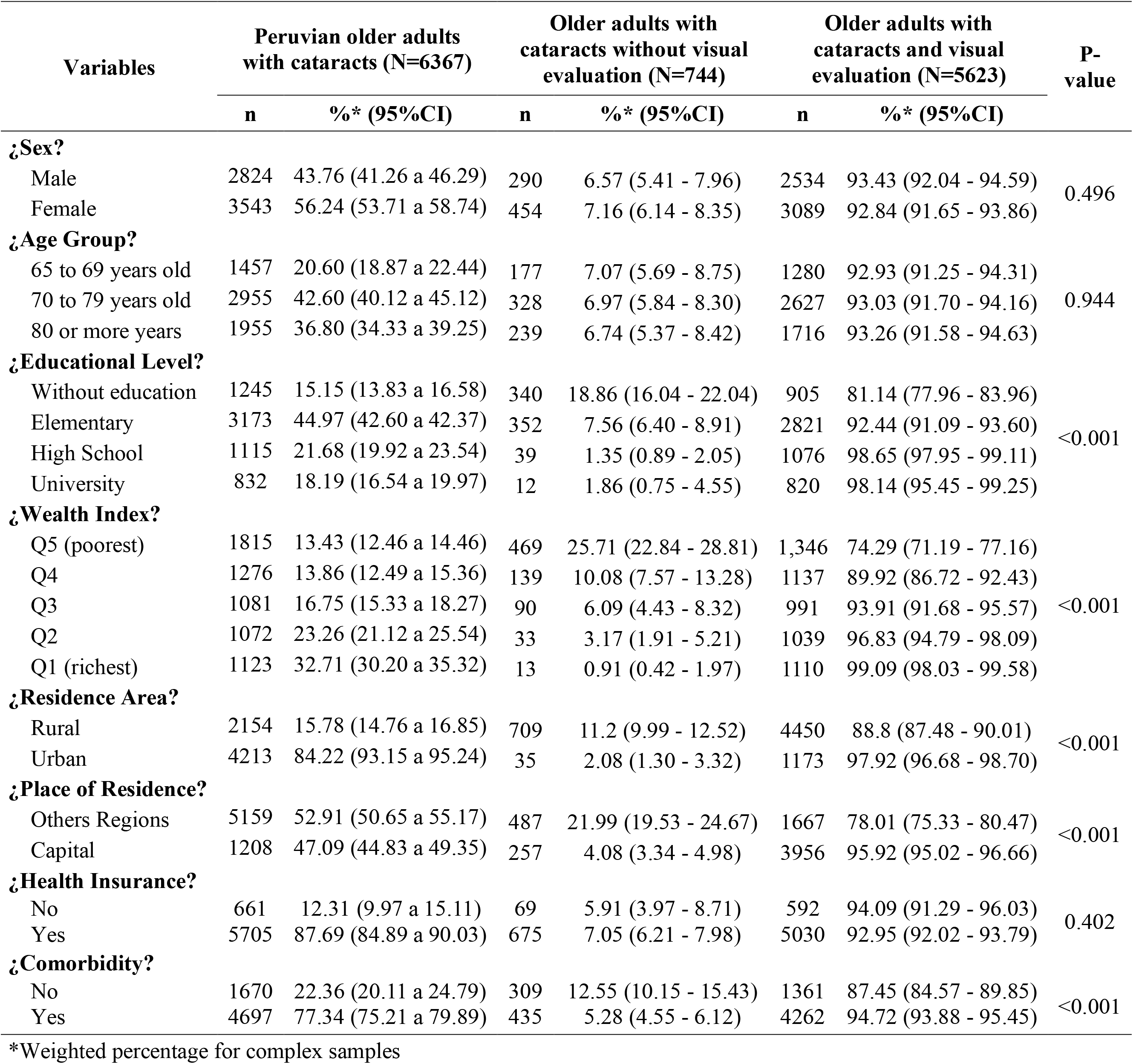
Sociodemographic characteristics of Peruvian older adults with cataracts according to visual evaluation.

In addition, characteristics such as level of education, wealth quintile, region or place of residence, and presence of any comorbidity were found to mediate differences in the proportion of adults with cataracts who had an eye examination (p*<*0.001). Specifically, older adults with cataract who had secondary education (aPR: 1.02; 95%CI: 1.00 to 1.04; p=0.047), who lived in the capital city (aPR: 1.02; 95%CI: 1.01 to 1.04; p=0.004), and who had any comorbidity (aPR: 1.04; 95%CI: 1.01 to 1.07; p=0.008) were more likely to have had an eye examination. While those with no education (aPR: 0.91; 95%CI: 0.87 to 0.94; p*<*0.001) or in the third (aPR: 0.97; 95%CI: 0.95 to 0.99; p=0.005), fourth (RPa: 0.95; 95%CI: 0.91 to 0.98; p=0.002), and fourth quintile (aPR: 0.95; 95%CI: 0.91 to 0.98; p=0.002) were more likely to have had an eye examination. and the last quintile (aPR: 0.82; 95%CI: 0.77 to 0.87; p*<*0.001) had a lower prevalence of an eye examination (Table 2).

**Table 2.** Regression model to assess sociodemographic characteristics of older adults with cataracts associated with visual evaluation.

| Variables | Crude Model |  |  | Adjusted Model |  |  |
| --- | --- | --- | --- | --- | --- | --- |
|  | cPR | 95%CI | P-value* | aPR | 95%CI | P-value* |
| **¿Sex?** |  |  |  |  |  |  |
| Male |  | **REF** |  |  | **REF** |  |
| Female | 0.994 | 0.976 - 1.012 | 0.493 | 0.995 | 0.978 - 1.012 | 0.541 |
| **¿Age Group?** |  |  |  |  |  |  |
| 65 to 69 years old |  | **REF** |  |  | **REF** |  |
| 70 to 79 years old | 0.997 | 0.977 - 1.018 | 0.810 | 0.997 | 0.978 - 1.017 | 0.780 |
| 80 or more years | 0.996 | 0.974 - 1.020 | 0.762 | 0.988 | 0.966 - 1.011 | 0.296 |
| **¿Educational Level?** |  |  |  |  |  |  |
| Without education |  | **REF** |  |  | **REF** |  |
| Elementary | 1.005 | 0.987 - 1.023 | 0.579 | 1.020 | 1.000 - 1.040 | 0.047 |
| High School | 0.942 | 0.922 - 0.963 | <0.001 | 0.994 | 0.971 - 1.017 | 0.583 |
| University | 0.827 | 0.794 - 0.861 | <0.001 | 0.906 | 0.872 - 0.941 | <0.001 |
| **¿Wealth Index?** |  |  |  |  |  |  |
| Q5 (poorest) |  | **REF** |  |  | **REF** |  |
| Q4 | 0.977 | 0.96 - 0.995 | 0.012 | 0.985 | 0.966 - 1.005 | 0.132 |
| Q3 | 0.948 | 0.927 - 0.968 | <0.001 | 0.969 | 0.948 - 0.991 | 0.005 |
| Q2 | 0.907 | 0.879 - 0.937 | <0.001 | 0.946 | 0.914 - 0.980 | 0.002 |
| Q1 (richest) | 0.750 | 0.72 - 0.781 | <0.001 | 0.818 | 0.772 - 0.866 | <0.001 |
| **¿Residence Area?** |  |  |  |  |  |  |
| Rural |  | **REF** |  |  | **REF** |  |
| Urban | 0.813 | 0.786 - 0.841 | <0.001 | 0.970 | 0.925 - 1.017 | 0.210 |
| **¿Place of Residence?** |  |  |  |  |  |  |
| Others Regions |  | **REF** |  |  | **REF** |  |
| Capital | 1.103 | 1.084 - 1.122 | <0.001 | 1.024 | 1.008 - 1.041 | 0.004 |
| **¿Health Insurance?** |  |  |  |  |  |  |
| No |  | **REF** |  |  | **REF** |  |
| Yes | 0.988 | 0.962 - 1.014 | 0.370 | 0.989 | 0.966 - 1.012 | 0.335 |
| **¿Comorbidity?** |  |  |  |  |  |  |
| No |  | **REF** |  |  | **REF** |  |
| Yes | 1.083 | 1.050 - 1.118 | <0.001 | 1.040 | 1.010 - 1.070 | 0.008 |

**cPR:** Crude Prevalence Ratio, **aPR;** Prevalence Ratio adjusted for the other variables, **95%CI:** 95% Confidence Interval, **REF:** Reference for estimating the measure of association in the other categories.
\*P-value estimated using the Poisson regression model

Regarding the inequality of ophthalmological assessment in older adults with cataracts, a moderate inequality was found in those without ophthalmological assessment (Figure 1A). In addition, adults with only primary education (CI: -0.20), living in rural areas (CI: -0.97) or in regions outside Lima (CI: -0.41), and without health insurance (CI: -0.22) were more unequal in having an ophthalmological examination. On the other hand, those with higher education (CI: 0.35) and those living in Lima (CI: 0.03) had less inequality in visual examinations (Figure 2).

**Figure 1.**
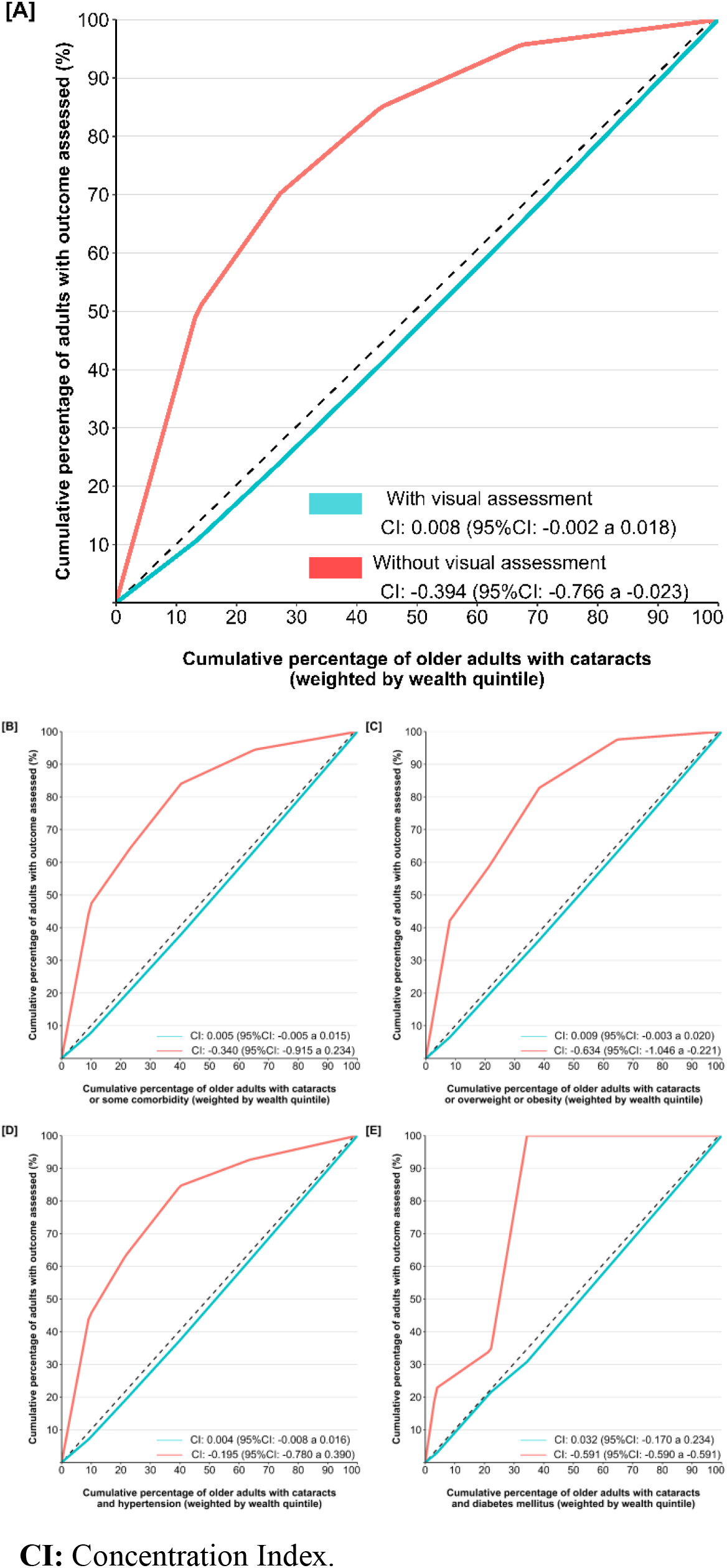
Inequality concentration index for visual assessment of older Peruvian adults with cataract according to comorbidities

**Figure 2.**
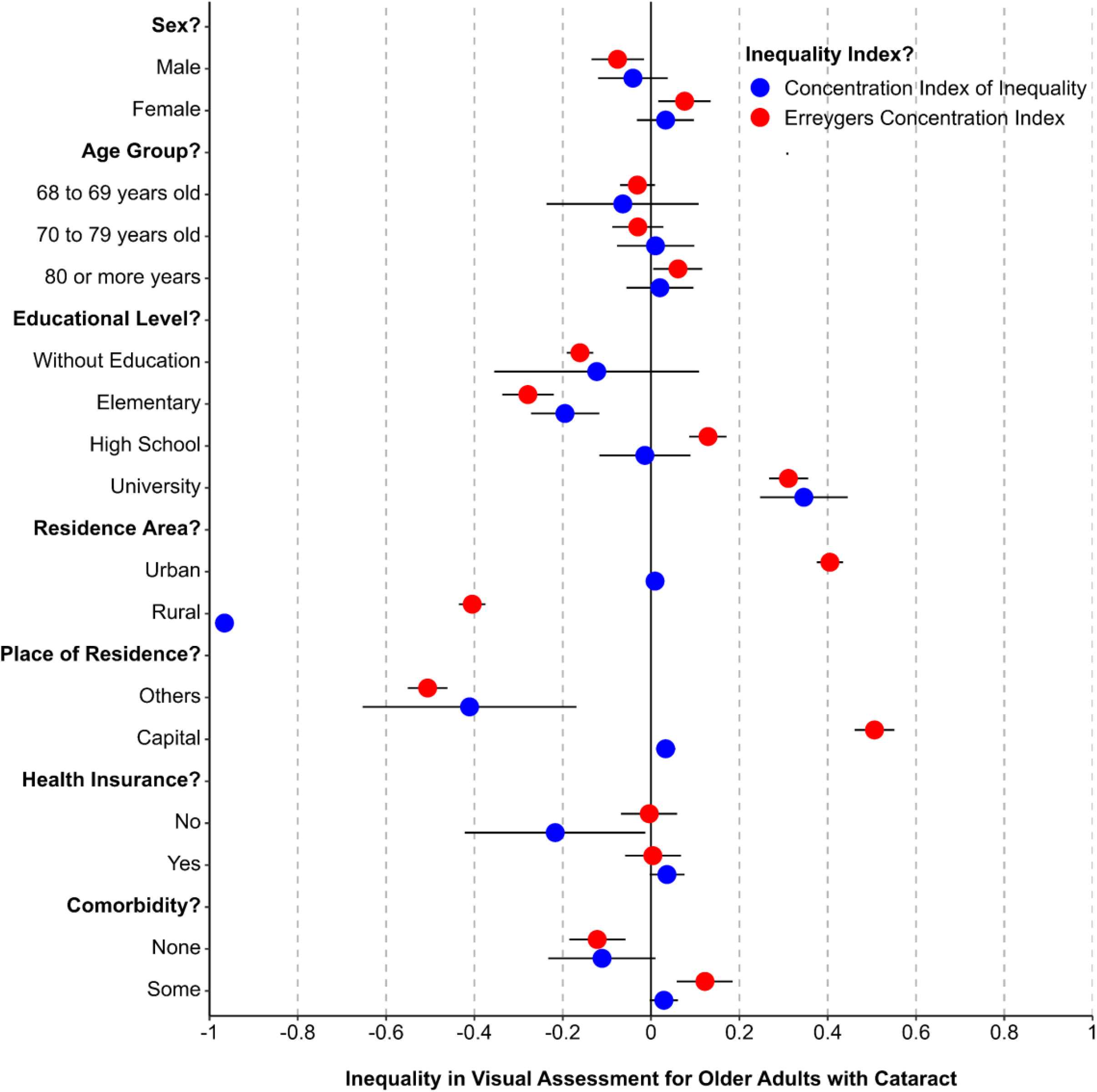
Inequality for visual assessment of older Peruvian adults with cataract according to socioeconomic conditions

Moreover, older adults with cataracts but no other health conditions showed higher inequality in receiving eye examinations (CI: -0.11 and ECI: -0.12). While inequality was higher among adults with cataracts and no visual assessment, it was higher among those with other comorbidities such as overweight or obesity (CI: -0.63) or diabetes mellitus (CI: -0.59), as shown in Figure 1B. Also, the proportion of older adults with cataracts increased from 2013 (24.8; 95%CI: 19.6 to 31.0) to 2023 (26.4; 95%CI: 23.7 to 29.3). The percentage of people who had an eye exam stayed the same between 19.6% (95%CI: 17.2 to 22.0) and 27.0% (95%CI: 23.8 to 30.1), but there was less inequality in eye exams from 2013 (ECI: 0.15) to 2020 (ECI: 0.07) and even more in 2023 (ECI: 0.12), as shown in Figure 3.

**Figure 3.**
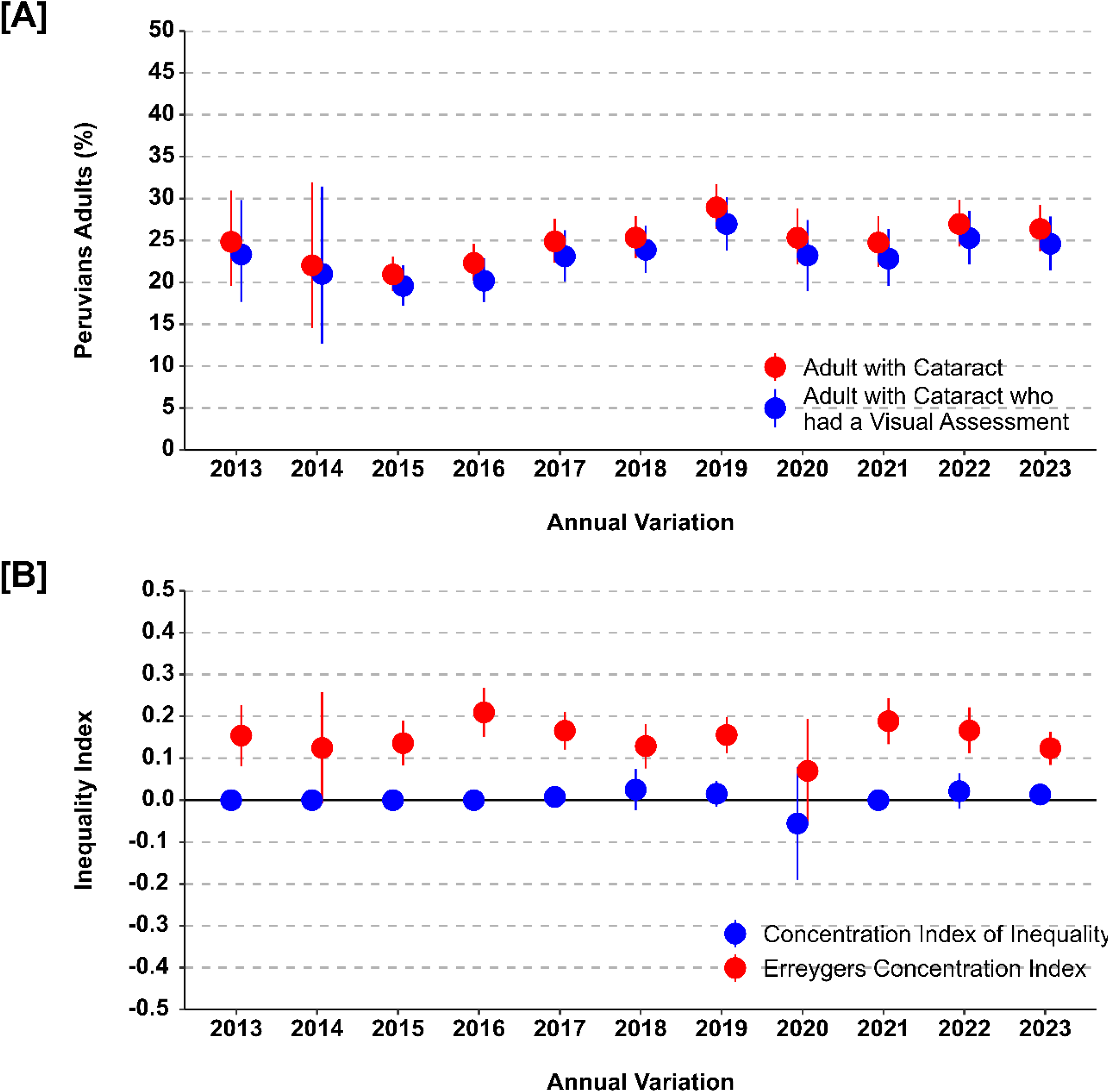
Annual variation of inequality for visual assessment of older adults with cataract

On the other hand, in terms of geographical distribution at the national level, a considerable variation in older adults with cataracts was observed in regions such as Cajamarca (13.8%) and Arequipa (32.4%). A lot of adults in this group had their eyes checked, but the coverage varied a lot between places like Huancavelica (68.3% of the population) and the constitutional province of Callao (96.9%). There were also big differences in the visual assessment for older adults with cataracts was spread out across the country (Figure 4). For instance, variations were observed between regions like Tumbes (ECI: 0.06) or Cusco (ECI: 0.29) and Ancash (ECI: 0.29).

**Figure 4.**
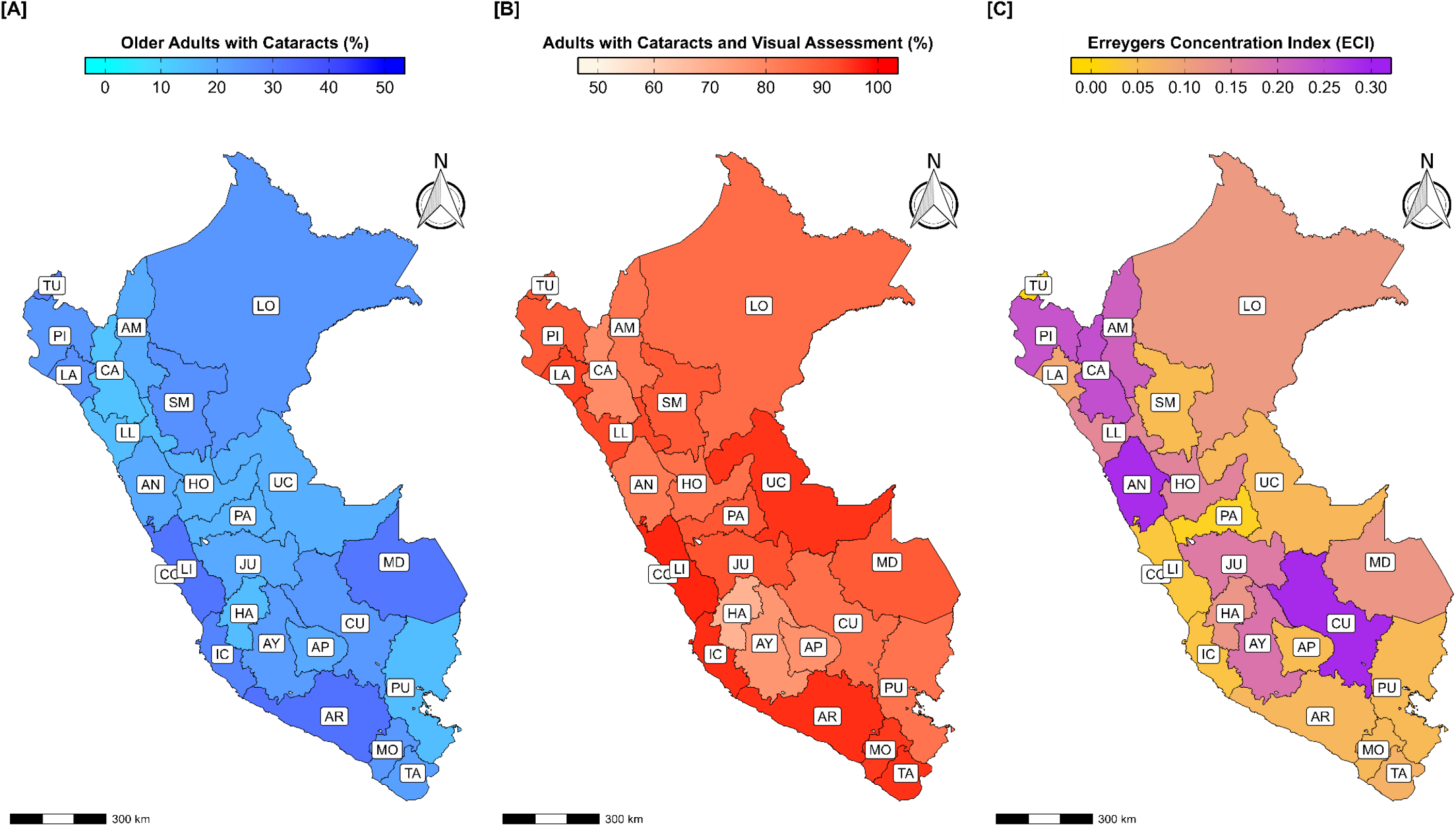
Geographical distribution of inequality for visual assessment of older Peruvian adults with cataract

## DISCUSSION

In this study, we evaluated inequalities in the visual assessment of older Peruvian adults with cataracts from 2013 to 2023. We found that those older adults living in rural areas experience greater disparities in receiving visual assessments, a gap that only increases when seeking surgery for cataract detection (35). Challenges such as limited access to healthcare facilities, irregular screening practices, and financial barriers worsen the disparities in timely diagnosis and treatment for eye conditions (36-40). In addition, the scarcity of ophthalmologists in Peru, with only 39.6 per million inhabitants, compared to countries like Argentina (103.6), Brazil (67.4), and Chile (49.8), perpetuates these inequalities (41,42).

Inequalities in the ophthalmological assessment of older adults with cataracts may be due to differences in waiting times for surgery, which are longer for women and adults with low educational attainment (43-45). This is consistent with studies indicating that older adults from disadvantaged socioeconomic backgrounds have reduced access to essential eye care services, leading to lower utilization rates (46). Therefore, the absence of visual care in the community restricts the availability of vision screening for the population, especially when additional costs are incurred due to the absence of ophthalmological screening covered by national health insurance (47,48).

In 2019, 16.6% of adults aged 50 years and older in Peru reported a diagnosis of cataract, which is lower than the estimated figures for Peruvian older adults with cataract between 2013 and 2023 (49). This figure is worrying given that cataracts account for 58.0% and 59.3% of cases of blindness and severe visual impairment, respectively, in Peru (40). As a result, older adults with limited education experience greater inequalities in visual assessment due to lower rates of visual examinations and follow-up care for cataract detection (50). This represent also an opportunity for educational interventions for visual health in older adults.

Moreover, older adults with cataracts and other health conditions exhibited fewer disparities in their visual assessments. This is notable given the higher likelihood of cataract development in individuals with diabetes mellitus and arterial hypertension (51). Furthermore, as the incidence of these chronic diseases increases with age, in Peru, between 20% and 35% of adults with diabetes had cataracts (52–54). Thus, routine visual assessment of patients with metabolic diseases is crucial from the first level of healthcare (55,56). Routine evaluations could prevent the development of cataracts or others visual diseases.

On the other hand, the COVID-19 pandemic reduced ophthalmological care and the number of elective cataract surgeries (57,58). In Peru, for instance, the impact of the pandemic increased inequalities in eye care (59). Despite the containment of the disease spread, the stagnation of eye health indicators at levels below those before the pandemic poses ongoing challenges for eye care in Peru (60). It is therefore crucial to develop strategies to expand screening, diagnosis, and treatment of eye health problems, as in Peru, the guidelines for managing this disease do not consider the inequalities faced by the population (25,26).

However, there were limitations because the ENDES does not address more aspects related to the development of cataracts in older adults. In addition, the social desirability bias could mediate an overestimation in the number of adults with visual assessments. The years in which the COVID-19 pandemic affected Peru could also modify the precision of the estimates because the form of data collection in the ENDES was modified to virtual platforms. Furthermore, it is unclear what level of understanding these adults have regarding the significance of visual evaluation, their practices for preventing eye health issues, and their responses when dealing with cataracts.

In conclusion, older adults with cataracts living in rural areas or outside Lima and having a lower educational level experienced greater disparities in vision assessments. These results underscore the critical importance of rectifying these disparities to guarantee appropriate eye care and advocate for tailored strategies to enhance the equity of vision screening among vulnerable populations, like older adults. It is vital to tackle these discrepancies in accessing eye care services to guarantee that every older adult, irrespective of their socioeconomic status or location, has equitable access to vision assessments and treatment.

## Data Availability

The data obtained from the National Institute of Statistics and Informatics (or INEI, in Spanish) platform: https://proyectos.inei.gob.pe/microdatos/

## Conflict of interests

None.

## Funding

Self-funded

## Acknowledgements

We thank the National Institute of Neoplastic Diseases in Peru for the development of the Demographic and Family Health Survey.

## Notes

### Competing Interest Statement

The authors have declared no competing interest.

### Funding Statement

This study did not receive any funding

### Author Declarations

The data was obtained from the National Institute of Statistics and Informatics (or INEI, in Spanish) platform: https://proyectos.inei.gob.pe/microdatos/

## REFERENCES

1. Cicinelli MV, Buchan JC, Nicholson M, Varadaraj V, Khanna RC. Cataracts. Lancet. 2023 Feb 4;401(10374):377–389. doi: 10.1016/S0140-6736(22)01839-6

2. GBD 2019 Blindness and Vision Impairment Collaborators; Vision Loss Expert Group of the Global Burden of Disease Study. Causes of blindness and vision impairment in 2020 and trends over 30 years, and prevalence of avoidable blindness in relation to VISION 2020: the Right to Sight: an analysis for the Global Burden of Disease Study. Lancet Glob Health. 2021;9(2):e144–e160. doi: 10.1016/S2214-109X(20)30489-7

3. Burton MJ, Ramke J, Marques AP, et al. The Lancet Global Health Commission on Global Eye Health: visión beyond 2020. Lancet Glob Health. 2021;9(4):e489–e551. doi: 10.1016/S2214-109X(20)30488-5

4. Mencucci R, Stefanini S, Favuzza E, Cennamo M, De Vitto C, Mossello E. Beyond vision:Cataract and health status in old age, a narrative review. Front Med (Lausanne). 2023;10:1110383. doi: 10.3389/fmed.2023.1110383

5. Lange N, Kujawska-Danecka H, Wyszomirski A, et al. Significant improvements in cataract treatment and persistent inequalities in access to cataract surgery among older Poles from 2009 to 2019: results of the PolSenior and PolSenior2 surveys. Front Public Health. 2023;11:1201689. doi: 10.3389/fpubh.2023.1201689

6. Emamian MH, Zeraati H, Majdzadeh R, Shariati M, Hashemi H, Fotouhi A. Economic inequality in eye care utilization and its determinants: a Blinder-Oaxaca decomposition. Int J Health Policy Manag. 2014;3(6):307–13. doi: 10.15171/ijhpm.2014.100

7. Trimmel J. Inequality and inequity in eye health. Community Eye Health. 2016;29(93):1–3. Available from: https://bit.ly/3VsVXzf

8. Buscho SE, Sharifi A, Cayenne S, et al. Racial Disparities in Cataract Surgery Timeline and Intraocular Lens Selection: A Retrospective Study. Transl Vis Sci Technol. 2023;12(11):20. doi: 10.1167/tvst.12.11.20

9. Su NH, Moxon NR, Wang A, French DD. Associations of Social Determinants of Health and Self-Reported Visual Difficulty: Analysis of the 2016 National Health Interview Survey. Ophthalmic Epidemiol. 2020;27(2):93–97. doi: 10.1080/09286586.2019.1680703

10. Lin X, Lou L, Miao Q, et al. The pattern and gender disparity in global burden of age-related macular degeneration. Eur J Ophthalmol. 2021;31(3):1161–1170. doi: 10.1177/1120672120927256

11. Williams A, Sahel J. Addressing Social Determinants of Vision Health. Ophthalmol Ther. 2022;11:1371–1382. doi: 10.1007/s40123-022-00531-w

12. Fang R, Yu YF, Li EJ, Lv NX, Liu ZC, Zhou HG, Song XD. Global, regional, national burden and gender disparity of cataract: findings from the global burden of disease study 2019. BMC Public Health. 2022;22(1):2068. doi: 10.1186/s12889-022-14491-0

13. Andean Health Organization-Hipolito Unanue Agreement. Situational Diagnosis of Eye Health by Life Course in the Andean Countries. 2022. Available from: https://bit.ly/3Vww4gX

14. Limburg H, Silva JC, Foster A. Cataract in Latin America: findings from nine recent surveys. Rev Panam Salud Publica. 2009;25(5):449–55. doi: 10.1590/s1020-49892009000500011

15. World Health Organization. World vision report. 2020. Available from: https://bit.ly/4bMsGFv

16. Bendezu-Quispe G, Fernandez-Guzman D, Caira-Chuquineyra B et al. Factors associated with the presence of cataracts in the Peruvian population older than 50 years: a cross-sectional study. F1000Research. 2022, 11:688. doi: 10.12688/f1000research.121802.1

17. Liu YC, Wilkins M, Kim T, Malyugin B, Mehta JS. Cataracts. Lancet. 2017;390(10094):600–612. doi: 10.1016/S0140-6736(17)30544-5

18. Jiang C, Melles RB, Sangani P, et al. Association of Behavioral and Clinical Risk Factors With Cataract: A Two-Sample Mendelian Randomization Study. Invest Ophthalmol Vis Sci. 2023;64(10):19. doi: 10.1167/iovs.64.10.19

19. Elam AR, Tseng VL, Rodriguez TM, et al. Disparities in Vision Health and Eye Care. Ophthalmology. 2022;129(10):e89–e113. doi: 10.1016/j.ophtha.2022.07.010

20. Li L, Wan X, Zhao G. Meta-analysis of the risk of cataract in type 2 diabetes. BMC Ophthalmol. 2014;14(94). doi: 10.1186/1471-2415-14-94

21. Kiziltoprak H, Tekin K, Inanc M, Goker YS. Cataract in diabetes mellitus. World J Diabetes. 2019;10(3):140–153. doi: 10.4239/wjd.v10.i3.140

22. Vedachalam R, Yamini K, Venkatesh R, et al. Reasons for delay in cataract surgery in patients with advanced cataracts during the COVID-19 pandemic. Indian J Ophthalmol. 2022;70(6):2153–2157. doi: 10.4103/ijo.IJO-544-22

23. Sebabi FO, Okello WO, Nakubulwa F, et al. Factors associated with delayed uptake of cataract surgery among adult patients at Mulago National Referral Hospital, Uganda. Afr Health Sci. 2021;21(3):1259–1265. doi: 10.4314/ahs.v21i3.36

24. López E, Álvarez-Dardet C, Gil-GonzÁlez D. Scientific Evidence and Recommendations on Vision Screening. Rev Esp Salud Pública. 2012; 86: 575–588. Available from: https://bit.ly/3XcwfR0

25. Ministry of Health. Technical Document: Clinical Practice Guidelines for Screening, Detection and Treatment of Cataract. Ministerial Resolution No. 537-2009-MINSA. 2009. Available from: https://bit.ly/3VdjDWZ

26. Ministry of Health. Technical Document: Management Plan for Eye Health and Prevention of Blindness. Ministerial Resolution No. 734-2022-MINSA. 2022. Available from: https://bit.ly/3x7uQQR

27. Yan W, Wang W, van Wijngaarden P, Mueller A, He M. Longitudinal changes in global cataract surgery rate inequality and associations with socioeconomic indices. Clin Exp Ophthalmol. 2019;47(4):453–460. doi: 10.1111/ceo.13430

28. Alinia C, Mohammadi SF, Jabbarvand M, Hashemi H. Geographical inequality in cataract surgery among Iranians between 2006 and 2011. East Mediterr Health J. 2018;24(7):664–671. doi: 10.26719/2018.24.7.664

29. Kyei S, Amponsah BK, Asiedu K, Akoto YO. Visual function, spectacle independence, and patients’ satisfaction after cataract surgerya study in the Central Region of Ghana. Afr Health Sci. 2021;21(1):445–456. doi: 10.4314/ahs.v21i1.55

30. Kheirkhah F, Roustaei G, Mohebbi E, et al. Improvement in Cognitive Status and Depressive Symptoms Three Months after Cataract Surgery. Caspian J Intern Med. 2018;9(4):386–392. doi: 10.22088/cjim.9.4.386

31. Xiao G, Zhu Z, Xiao X, et al. Geographical Inequality on Cataract Surgery Uptake in 200,000 Australians: Findings from the “45 and Up Study”. Comput Intell Neurosci. 2022;2022:9618912. doi: 10.1155/2022/9618912

32. National Institute of Statistics and Informatics. Peru: Population Estimates and Projections by Department, Sex and Five-Year Age Groups 1995-2025. Demographic Analysis Bulletin N°37. 2009. Available from: https://bit.ly/4caY2p9

33. O’Donnell O, O’Neill S, Van Ourti T, Walsh B. conindex: Estimation of concentration indices. Stata J. 2016 1st Quarter;16(1):112–138. Available from: https://bit.ly/3KwlMYT

34. Erreygers G. Correcting the Concentration Index. J Health Econ. 2009;28(2):504–15. doi: 10.1016/j.jhealeco.2008.02.003

35. An L, Jan CL, Feng J, et al. Inequity in Access: Cataract Surgery Throughput of Chinese Ophthalmologists from the China National Eye Care Capacity and Resource Survey. Ophthalmic Epidemiol. 2020;27(1):29–38. doi: 10.1080/09286586.2019.1678654

36. Wu AM, Wu CM, Tseng VL, et al. Characteristics Associated With Receiving Cataract Surgery in the US Medicare and Veterans Health Administration Populations. JAMA Ophthalmol. 2018;136(7):738–745. doi: 10.1001/jamaophthalmol.2018.1361

37. Kauh CY, Blachley TS, Lichter PR, et al. Geographic Variation in the Rate and Timing of Cataract Surgery Among US Communities. JAMA Ophthalmol. 2016; 134(3):267. doi: 10.1001/jamaophthalmol.2015.5322

38. Lee CS, Su GL, Baughman DM, Wu Y, Lee AY (2017) Disparities in delivery of ophthalmic care; An exploration of public Medicare data. PLoS ONE. 12(8): e0182598. doi: 10.1371/journal.pone.0182598

39. Kilmer G, Bynum L, Balamurugan A. Access to and use of eye care services in rural arkansas. J Rural Health. 2010 Winter;26(1):30–5. doi: 10.1111/j.1748-0361.2009.00262.x

40. Patel D, Ananthakrishnan A, Lin T, et al. Social Determinants of Health and Impact on Screening, Prevalence, and Management of Diabetic Retinopathy in Adults: A Narrative Review. J. Clin. Med. 2022, 11, 7120. doi: 10.3390/jcm11237120

41. Resnikoff S, Lansingh VC, Washburn L, et al. Br J Ophthalmol. 2020;104:588–592

42. Balas M, Vasiliu D, Austria G, et al. Demographic trends of patients undergoing ophthalmic surgery in Ontario, Canada: a population-based study. BMJ Open Ophthalmology. 2023;8:e001253. doi:10.1136/bmjophth-2023-001253

43. Smirthwaite G, Lundström M, Wijma B, et al. Inequity in waiting for cataract surgery–an analysis of data from the Swedish National Cataract Register. Int J Equity Health. 2016;15:10. doi: 10.1186/s12939-016-0302-3

44. Tielsch JM, Sommer A, Witt K, Katz J, Royall RM. Blindness and visual impairment in an American urban population. The Baltimore Eye Survey. Arch Ophthalmol. 1990;108(2):286–90. doi: 10.1001/ar-chopht.1990.01070040138048

45. Desai N, Copeland RA. Socioeconomic disparities in cataract surgery. Curr Opin Ophthalmol. 2013;24(1):74–8. doi: 10.1097/ICU.0b013e32835a93da

46. Matta S, Park J, Palamaner G, et al. Cataract Surgery Visual Outcomes and Associated Risk Factors in Secondary Level Eye Care Centers of L V Prasad Eye Institute, India. PLoS ONE. 2016;11(1):e0144853. doi:10.1371/journal.pone.014485

47. Vinluan ML, Olveda RM, Olveda DU, et al. BMJ Case Rep Published online: [ please include Day Month Year] doi:10.1136/bcr-2014-208197

48. Zhang X, Cotch MF, Ryskulova A, et al. Vision health disparities in the United States by race/ethnicity, education, and economic status: findings from two nationally representative surveys. Am J Ophthalmol. 2012;154(6 Suppl):S53-62.e1. doi: 10.1016/j.ajo.2011.08.045

49. Campos B, Cerrate A, Montjoy E, et al. National survey on the prevalence and causes of blindness in Peru. Rev Panam Salud Publica. 2014;36(5):283–9. Available from: https://bit.ly/4caY6VV

50. Drinkwater JJ, Davis WA, Davis TME. A systematic review of risk factors for cataract in type 2 diabetes. Diabetes Metab Res Rev. 2019;35(1):e3073. doi: 10.1002/dmrr.3073

51. Srinivasan S, Raman R, Swaminathan G, et al. Incidence, Progression, and Risk Factors for Cataract in Type 2 Diabetes. Invest Ophthalmol Vis Sci. 2017;58(13):5921–5929. doi: 10.1167/iovs.17-22264

52. Pék A, Szabó D, SÁndor GL, et al. Relationship between diabetes mellitus and cataract in Hungary. Int J Ophthalmol. 2020;13(5):788–93.

53. Behera UC, Salzman B, Das AV, et al. Prevalence of chronic disease in older adults in multitier eye-care facilities in South India: Electronic medical records-driven big data analytics report. Indian J Ophthalmol. 2021;69(12):3618–3622. doi: 10.4103/ijo.IJO62121

54. Webb EM, Rheeder P, Roux P. Screening in Primary Care for Diabetic Retinopathy, Maculopathy and Visual Loss in South Africa. Ophthalmologica. 2016;235(3):141–9. doi: 10.1159/000443972

55. Ammary-Risch NJ, Huang SS. The primary care physician’s role in preventing vision loss and blindness in patients with diabetes. J Natl Med Assoc. 2011;103(3):281–3. doi: 10.1016/s0027-9684(15)30288-1

56. Cetinkaya YF. Ophthalmic surgeries before and during the covid-19 outbreak in a tertiary hospital. Int Ophthalmol. 2023;43(5):1565–1570. doi: 10.1007/s10792-022-02555-4

57. Yen CY, Fang IM, Tang HF, et al. COVID-19 pandemic decreased the ophthalmic outpatient numbers and altered the diagnosis distribution in a community hospital in Taiwan: An observational study. PLoS One. 2022;17(3):e0264976. doi: 10.1371/journal.pone.0264976

58. Roberti J, Leslie HH, Doubova SV, et al. Inequalities in health system coverage and quality: a cross-sectional survey of four Latin American countries. Lancet Glob Health. 2024;12(1):e145–e155. doi: 10.1016/S2214-109X(23)00488-6

59. Ahmad K, Zwi AB, Tarantola DJ, et al. Gendered Disparities in Quality of Cataract Surgery in a Marginalised Population in Pakistan: The Karachi Marine Fishing Communities Eye and General Health Survey. PLoS One. 2015;10(7):e0131774. doi: 10.1371/journal.pone.0131774

60. Aghaji A, Burchett HED, Oguego N, et al. Primary health care facility readiness to implement primary eye care in Nigeria: equipment, infrastructure, service delivery and health management information systems. BMC Health Serv Res. 2021;21(1):1360. doi: 10.1186/s12913-021-07359-3

